# Multi-ancestral genome-wide association study of clinically defined nicotine dependence reveals strong genetic correlations with other substance use disorders and health-related traits

**DOI:** 10.1101/2025.01.29.25320962

**Authors:** Emma C Johnson, Dongbing Lai, Alex P Miller, Alexander S Hatoum, Joseph D Deak, Jared V Balbona, David AA Baranger, Marco Galimberti, Kittipong Sanichwankul, Thorgeir Thorgeirsson, Sarah MC Colbert, Sandra Sanchez-Roige, Keyrun Adhikari, Anna Docherty, Louisa Degenhardt, Tobias Edwards, Louis Fox, Alexandros Giannelis, Paul Jeffries, Tellervo Korhonen, Claire Morrison, Yaira Z Nunez, Teemu Palviainen, Mei-Hsin Su, Pamela N Romero Villela, Leah Wetherill, Emily A Willoughby, Stephanie Zellers, Laura Bierut, Jadwiga Buchwald, William Copeland, Robin Corley, Naomi P. Friedman, Tatiana M Foroud, Nathan A Gillespie, Ian R Gizer, Andrew C Heath, Ian B Hickie, Jaakko A Kaprio, Matthew C Keller, James L Lee, Penelope A Lind, Pamela A Madden, Hermine HM Maes, Nicholas G Martin, Matt McGue, Sarah E Medland, Elliot C Nelson, John V Pearson, Bernice Porjesz, Michael Stallings, Scott Vrieze, Kirk C Wilhelmsen, Raymond K Walters, Renato Polimanti, Robert T Malison, Hang Zhou, Kari Stefansson, Marc N Potenza, Apiwat Mutirangura, Vorasuk Shotelersuk, Rasmon Kalayasiri, Howard J Edenberg, Joel Gelernter, Arpana Agrawal

## Abstract

Genetic research on nicotine dependence has utilized multiple assessments that are in weak agreement. We conducted a genome-wide association study of nicotine dependence defined using the Diagnostic and Statistical Manual of Mental Disorders (DSM-NicDep) in 61,861 individuals (47,884 of European ancestry, 10,231 of African ancestry, 3,746 of East Asian ancestry) and compared the results to other nicotine-related phenotypes. We replicated the well-known association at the *CHRNA5* locus (lead SNP: rs147144681, p =1.27E-11 in European ancestry; lead SNP = rs2036527, p = 6.49e-13 in cross-ancestry analysis). DSM-NicDep showed strong positive genetic correlations with cannabis use disorder, opioid use disorder, problematic alcohol use, lung cancer, material deprivation, and several psychiatric disorders, and negative correlations with respiratory function and educational attainment. A polygenic score of DSM-NicDep predicted DSM-5 tobacco use disorder and 6 of 11 individual diagnostic criteria, but none of the Fagerström Test for Nicotine Dependence (FTND) items, in the independent NESARC-III sample. In genomic structural equation models, DSM-NicDep loaded more strongly on a previously identified factor of general addiction liability than did a “problematic tobacco use” factor (a combination of cigarettes per day and nicotine dependence defined by the FTND). Finally, DSM-NicDep was strongly genetically correlated with a GWAS of tobacco use disorder as defined in electronic health records, suggesting that combining the wide availability of diagnostic EHR data with nuanced criterion-level analyses of DSM tobacco use disorder may produce new insights into the genetics of this disorder.

## INTRODUCTION

Nicotine use and nicotine dependence (or tobacco use disorder) are influenced by both genetic liability and environmental factors^1,2^. Several different smoking-related phenotypes have been studied, including smoking initiation^3^, cigarettes per day^3^, the Fagerström Test for Nicotine Dependence (FTND)^4^, problematic tobacco use^5^, tobacco use disorder defined by codes from the International Classification of Diseases (ICD-TUD)^6^, and nicotine dependence^7^ defined by the Diagnostic and Statistical Manual of Mental Disorders (DSM-NicDep) (**Table 1**).

**Table 1.** Definitions of different smoking-related phenotypes discussed in this manuscript, the acronyms used to represent them, and the relevant GWASs used in analyses.

| Smoking phenotype | Shorthand name used in current manuscript | Phenotype definition | Relevant GWAS PubMed ID |
| --- | --- | --- | --- |
| Smoking initiation | SmkInit | Whether someone has ever been a regular smoker. | 36477530 |
| Cigarettes per day | CPD | Number of cigarettes smoked per day. | 36477530 |
| Fagerström Test for Nicotine Dependence | FTND | A six-item questionnaire assessing the quantity of cigarettes smoked and compulsion to use, resulting in an ordinal measure of nicotine dependence. | 33144568 |
| Tobacco use disorder | ICD-TUD | Binary diagnosis defined using International Classification of Disease (ICD) codes (ICD 10 code for TUD: F17.200). | 38632388 |
| Nicotine dependence | DSM-NicDep | Seven criteria designed to assess clinical features of dependence, including tolerance and problems caused by using cigarettes; at least 3 of the 7 criteria must be endorsed to reach a diagnosis of nicotine dependence. | NA |
| Problematic tobacco use | PTU | A combination of FTND and CPD GWAS, produced using MTAG. | 34750568 |

Genome-wide association studies (GWAS) of nicotine use behaviors have largely focused on phenotypes that are easily ascertained through self-report questionnaires (e.g., *when did you start smoking?*, *are you a current smoker?*, and *how many cigarettes do you smoke in a day?*). The largest genome-wide association studies (GWAS) of tobacco smoking behaviors to date identified 140 loci associated with cigarettes per day (CPD; N = 784,353)^3^.

GWAS of nicotine dependence phenotypes have generally been smaller. Some GWAS of nicotine dependence-related phenotypes have focused on data collected from short questionnaires such as the Fagerström Test for Nicotine Dependence (FTND^4^). The FTND is a 6-item questionnaire which includes cigarettes smoked per day as an ordinal indicator. The largest GWAS of FTND (defined as 0–3 for mild, 4–6 for moderate, and 7–10 for severe dependence) identified 5 loci^4^. Given the strong genome-wide genetic correlation between CPD and FTND (r_g_=0.95)^4^, these GWAS were combined into a single phenotype, problematic tobacco use (PTU), in another study^5^ using multi-trait analysis of GWAS (MTAG). A recent GWAS meta-analysis of ICD-TUD from electronic health records (EHR; see **Table 1**) reached nearly 900,000 samples and identified 88 loci^6^, all of which had been implicated in prior smoking-related GWAS. ICD-TUD showed modest genetic correlations with cigarettes per day (r_g_ = 0.44)^6^ and FTND (r_g_ = 0.63).

Our study was motivated by epidemiological and clinical data supporting nosological distinctions between FTND and ICD or DSM-based diagnoses, including some studies that suggest qualitative and quantitative differences in associations between DSM- and FTND-defined nicotine dependence and some psychopathology^8,9^. The FTND is brief, and thus easily and frequently collected. It has been especially prioritized in clinical trials of tobacco cessation^10,11^, likely because FTND scores correlate well with relapse and treatment response and the scale places a great deal of emphasis on physiological aspects of dependence (e.g., items related to tolerance and withdrawal^12^). On the other hand, both ICD- and DSM-based nicotine dependence include criteria related to physical and psychological (and social, in DSM-5) impairment due to nicotine/tobacco use, as well as behaviors directed at seeking and using nicotine to the exclusion of other activities. Neither FTND nor the ICD-TUD diagnostic classification maps perfectly to DSM-NicDep^13,14^.

While many large GWAS meta-analyses of substance use disorders have relied on cases defined using clinical criteria recommended by DSM or ICD classification, the few GWAS of DSM-NicDep to date have been relatively small^7^. We conducted a large meta-analysis of DSM-NicDep, combining data across 16 cohorts and multiple genetic ancestries. The largest analyses of psychiatric traits have focused on individuals whose genetics most closely resemble the European ancestry subset of the 1000 genomes project, so we assessed the genetic correlations between DSM-NicDep and other substance use phenotypes, psychiatric disorders, and related phenotypes in that subset. We also compared the correlations of the various nicotine dependence related measures (DSM-NicDep, ICD-TUD, and PTU) and other substance use disorders (cannabis use disorder, problematic alcohol use, and opioid use disorder) by fitting a previously published common factor model (the Addiction-risk-factor^15^) to the data using genomic structural equation modeling. Finally, we tested whether a polygenic score for DSM-NicDep was associated with DSM-5 tobacco use disorder and its 11 diagnostic criteria, and 4 of the 6 FTND criteria, in a large, independent sample, the National Epidemiologic Survey on Alcohol and Related Conditions-III (NESARC-III) cohort.

## METHODS

### Genome-wide association study of DSM-defined nicotine dependence (DSM-NicDep)

We performed meta-analyses of GWASs of DSM-defined nicotine dependence (hereafter referred to as “DSM-NicDep”) across a total of 18 cohorts, three of which included samples of multiple ancestries, using a sample size-weighted meta-analysis implemented in METAL^16^. We used nicotine-exposed controls where possible (see **Supplemental Materials** for more details on each cohort). Genetic ancestry similarity was inferred by comparing an individual’s genome to the genomes from global reference populations using statistical methods such as principal components analysis, although exact methods varied somewhat across cohorts. There were 16 cohorts with samples that were most genetically similar to European ancestry global reference populations, 4 with samples that were most genetically similar to African ancestry global reference populations, and one cohort whose participants were most genetically similar to East Asian ancestry global reference populations (hereafter referred to as European, African, or East Asian ancestries). All GWAS controlled for age, sex, and 10 principal components; more details, including cohort-specific covariates, are provided in **Supplemental Table 1**. Twelve cohorts provided summary statistics, while five cohorts provided individual genotype and phenotype data for analysis (**Supplemental Table 1**).

We used FUMA v1.5.2^17^ to identify independent, genome-wide significant risk loci, annotate variants, and perform gene-wise analyses via MAGMA^18^. We used the default FUMA parameters to define “independent significant SNPs” as those which reached genome-wide significance (p < 5e-8) and were independent of each other at r2 < 0.6, and “lead SNPs” as those SNPs that were independent of each other at r2 < 0.1. “Genomic risk loci” were defined by merging linkage disequilibrium (LD) blocks of independent significant SNPs within a 250 kb distance. We performed gene mapping in FUMA using positional mapping (based on ANNOVAR annotations), expression quantitative trait locus (eQTL) mapping (using GTEx V8^19^, CommonMind^20^, and Braineac^21^ data), and chromatin interaction mapping. We also performed the MAGMA gene, gene-set, and gene expression analyses (using GTEx V8 data).

We performed follow-up analyses described below (genetic correlations, genomic structural equation modeling, and polygenic scores) only in the European ancestry data, because the African and East Asian ancestry subsets were under-powered.

### Genetic correlations with other substance use disorders, psychiatric disorders, and other relevant phenotypes

We used linkage disequilibrium score regression (LDSC^22,23^) to estimate the SNP-heritability of DSM-NicDep and the genetic correlations between DSM-NicDep and other substance use phenotypes, using published GWASs of problematic alcohol use (PAU^24^), Fagerström Test for Nicotine Dependence (FTND^4^), ICD-based tobacco use disorder (ICD-TUD^6^), problematic tobacco use (PTU, a combination of FTND and cigarettes per day^15^), cannabis use disorder (CanUD^25^), opioid use disorder (OUD^26^), smoking initiation ^3^, smoking cessation ^3^, and cigarettes per day^3^. We also estimated genetic correlations between DSM-NicDep and other phenotypes including psychiatric disorders, behavioral traits, respiratory health, and socioeconomic status-related phenotypes. Details on the individual GWAS used in genetic correlation analyses are provided in the **Supplemental Methods**. We further tested whether genetic correlations for DSM-NicDep and FTND were different from each other using a block-jackknife method^23,27^.

### Genomic structural equation modeling

We applied confirmatory factor analysis to the covariance matrix generated by LDSC using genomic structural equation modeling (genomic SEM^28^) with weighted least squares estimation. As in Hatoum et al.^5^, the indicators were allowed to load freely on a single latent factor (*Addiction-Risk-Factor*), but updated the OUD^26^, PAU^24^, and CanUD^25^ GWAS. We compared *Addiction-Risk-Factor* models with DSM-NicDep, PTU, or ICD-TUD as the tobacco-related indicator. The variance of the common latent factor was scaled to 1.0.

### Polygenic score analyses

We created polygenic scores for DSM-NicDep and FTND in the European ancestry subset of the NESARC-III sample. NESARC-III was genotyped using the Affymetrix Axiom® Exome Array^29^, which limited our ability to impute SNPs due to lack of appreciable non-exonic coverage and resulted in some regions with low SNP densities. Details of QC and imputation are available in the **Supplemental Materials.** We used PRS-CS^30^, a Bayesian method that uses continuous shrinkage priors to weight SNP effect sizes, and used the ‘auto’ function, which allows the global scaling parameter to be automatically learned from the data. We then used the ‘score’ function in Plink 1.9^31^ to create polygenic scores for DSM-NicDep and FTND in NESARC-III. The associations between DSM-5 tobacco use disorder, endorsement of the 11 individual diagnostic criteria, and 4 of the 6 FTND criteria (excluding cigarettes per day and smoking when ill) in NESARC-III (N = 12,482; N_cases_ = 4,205) and the polygenic scores for DSM-NicDep and FTND were estimated using logistic regression models in R, covarying for age, sex, and 10 within-ancestry principal components. Both polygenic scores were included jointly as predictors in the same model.

## RESULTS

### Genome-wide association study of DSM-defined nicotine dependence

There were a total of 15 cohorts included in the European ancestry (EUR) meta-analysis (N_cases_ = 20,923, N_controls_ = 26,961), four cohorts included in the African ancestry (AFR) meta-analysis (N_cases_ = 5,293, N_controls_ = 4,938), and one cohort included in the East Asian ancestry (EAS) GWAS (N_cases_ = 2,007, N_controls_ = 1,739), for a total cross-ancestry sample size of N = 61,861 (28,223 cases; **Supplemental Table 1**). We identified one genome-wide significant locus, near the cholinergic receptor nicotinic alpha 5 subunit (*CHRNA5*) gene on chromosome 15 (lead SNP: rs147144681, p-value =1.27E-11 in European ancestry; lead SNP = rs2036527, p-value = 6.49e-13 in cross-ancestry analysis; **Supplemental Tables 2 & 3**). Using a lifetime prevalence of 24%^32^, we estimated the liability scale SNP-heritability of DSM-defined nicotine dependence (DSM-NicDep) to be 0.07 (SE = 0.01) in the European ancestry samples.

In the MAGMA gene-based analysis of the European ancestry data, three genes were significant after multiple testing corrections: *CHRNA5, CHRNA3,* and *IREB2* (**Supplemental Table 4**). Three gene-sets were significantly associated: “GOBP_TRANSCRIPTION_BY_RNA_POLYMERASE_III”, “GOMF_PRE_MRNA_5_SPLICE_SITE_BINDING”, and “REACTOME_HIGHLY_CALCIUM_PERMEABLE_NICOTINIC_ACETYLCHOLINE_RECEPTO RS”. None of the 30 tissue types from GTEx v8 were significant.

### Genetic correlations with substance use and other phenotypes

In the EUR data, DSM-NicDep was strongly correlated with the other measures of nicotine dependence (r_g_s with FTND, PTU, and ICD-TUD ranged from 0.77 to 1.01). The genetic correlations between DSM-NicDep and published GWASs of substance use disorders were of moderate-to-high magnitude: cannabis use disorder (CanUD), problematic alcohol use (PAU), and opioid use disorder (OUD; r_g_ = 0.64 – 0.84). Overall, DSM-NicDep was significantly correlated with 23 of the 26 phenotypes tested (**Figure 1; Supplemental Table 5**). Compared to the other tobacco-related phenotypes, DSM-NicDep showed the strongest correlations with many traits, albeit with wider confidence intervals due to smaller sample size. When correcting for 24 comparisons, the genetic correlations between DSM-NicDep and smoking initiation, cannabis ever-use, CanUD, and the Townsend deprivation index were significantly larger (p < 0.002) than the corresponding genetic correlations between FTND and these phenotypes.

**Figure 1.**
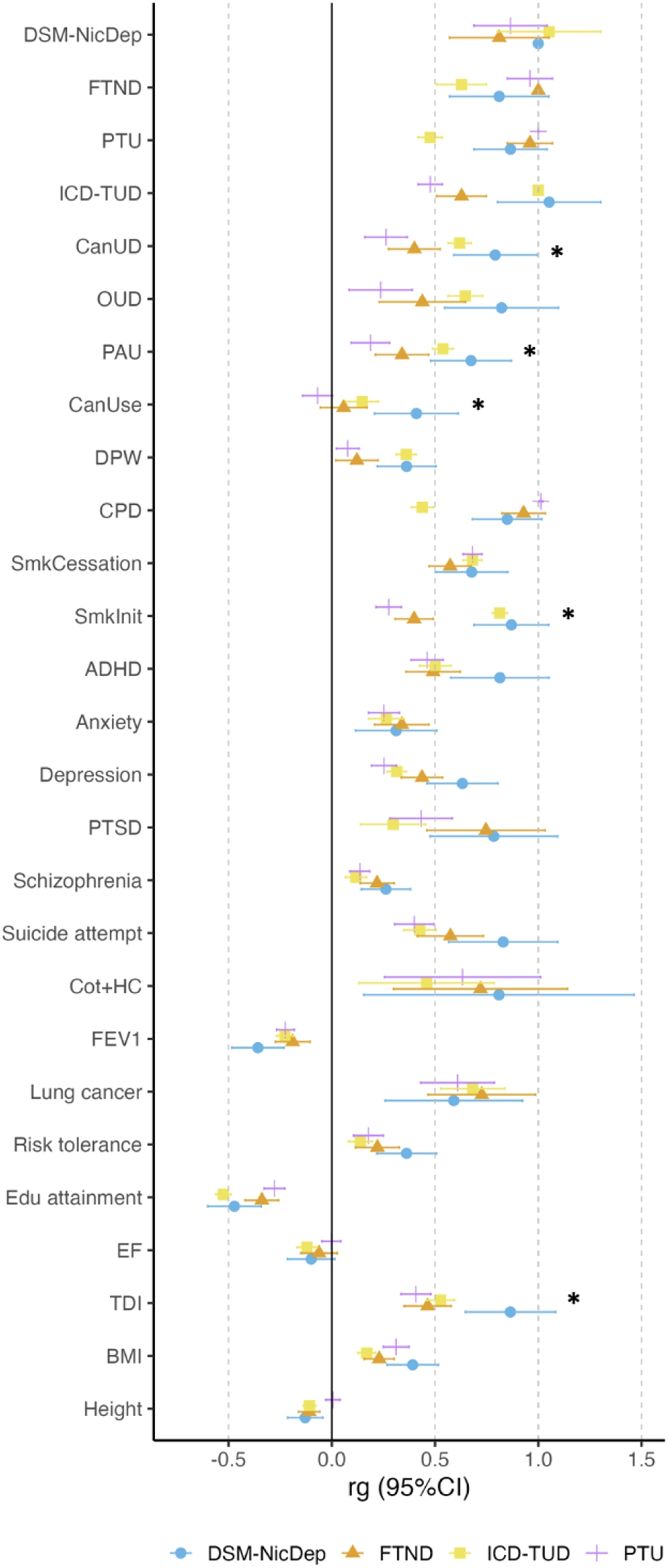
Comparing genetic correlations (r_g_) for DSM-NicDep, FTND, ICD-TUD, and PTU with other traits in European ancestry data. Traits include other substance use disorders (CanUD = cannabis use disorder^25^; OUD = opioid use disorder^26^; PAU = problematic alcohol use^24^, ICD-TUD = ICD-based tobacco use disorder^6^), substance use behaviors (CanUse = cannabis ever-use^33^; DPW = drinks per week^3^; SmkInit = smoking initiation^3^, SmkCessation = smoking cessation^3^, CPD = cigarettes per day^3^), psychiatric disorders and other mental health phenotypes (ADHD = attention deficit hyperactivity disorder^34^; PTSD = post-traumatic stress disorder^35^), biomarkers (Cot+HC = cotinine + 3-hydroxycotinine^36^), lung health-related traits (FEV1 = forced expiratory volume in 1 second), risk tolerance^37^, socioeconomic status-related traits (Edu attainment = educational attainment^38^; TDI = Townsend deprivation index), executive function (EF^39^), and anthropometric measures (BMI = body mass index^40^; height^41^). * indicates r_g_s that significantly differ between DSM-NicDep and FTND at = 0.002 (Bonferroni correction for 24 comparisons).

### Genomic structural equation models of broad addiction liability

In the EUR data, we found that a common genetic factor model with DSM-NicDep, PAU, OUD, and CanUD as indicators, similar to the *Addiction-Risk-Factor* presented in Hatoum et al.^4^, fit the data well (**Figure 2A; Supplemental Table 6**). There were two differences between the Hatoum et al. *Addiction-Risk-Factor* and our modified model: (1) we used larger, more recent versions of the OUD^26^, PAU^24^, and CanUD^25^ GWASs; and (2) we removed the residual correlation between PAU and OUD, as this path was no longer significant. We compared the model with DSM-NicDep as the tobacco-related indicator (**Figure 2A**) to one where DSM-NicDep was substituted by the original problematic tobacco use (PTU) GWAS meta-analysis (FTND + CPD) from Hatoum et al.^4^ (**Figure 2B; Supplemental Table 7**), and one where NicDep was substituted by ICD-TUD^6^ (**Figure 2C; Supplemental Table 8**). Each of the modified models fit the data well, but the loading for DSM-NicDep was the largest of the three tobacco-related indicators, almost 3-fold larger than the loading for PTU in the equivalent model (0.86 vs. 0.30).

**Figure 2.**
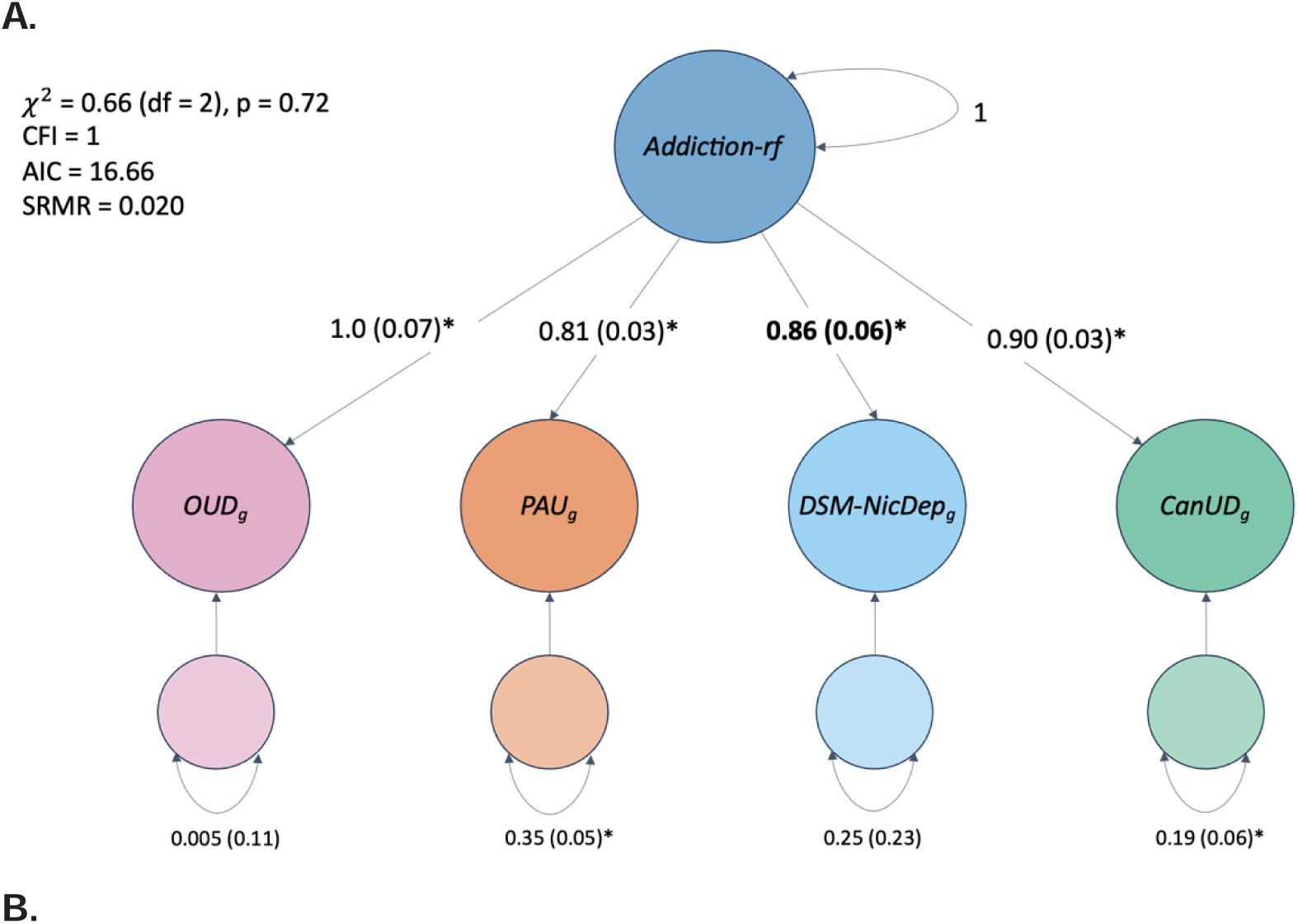
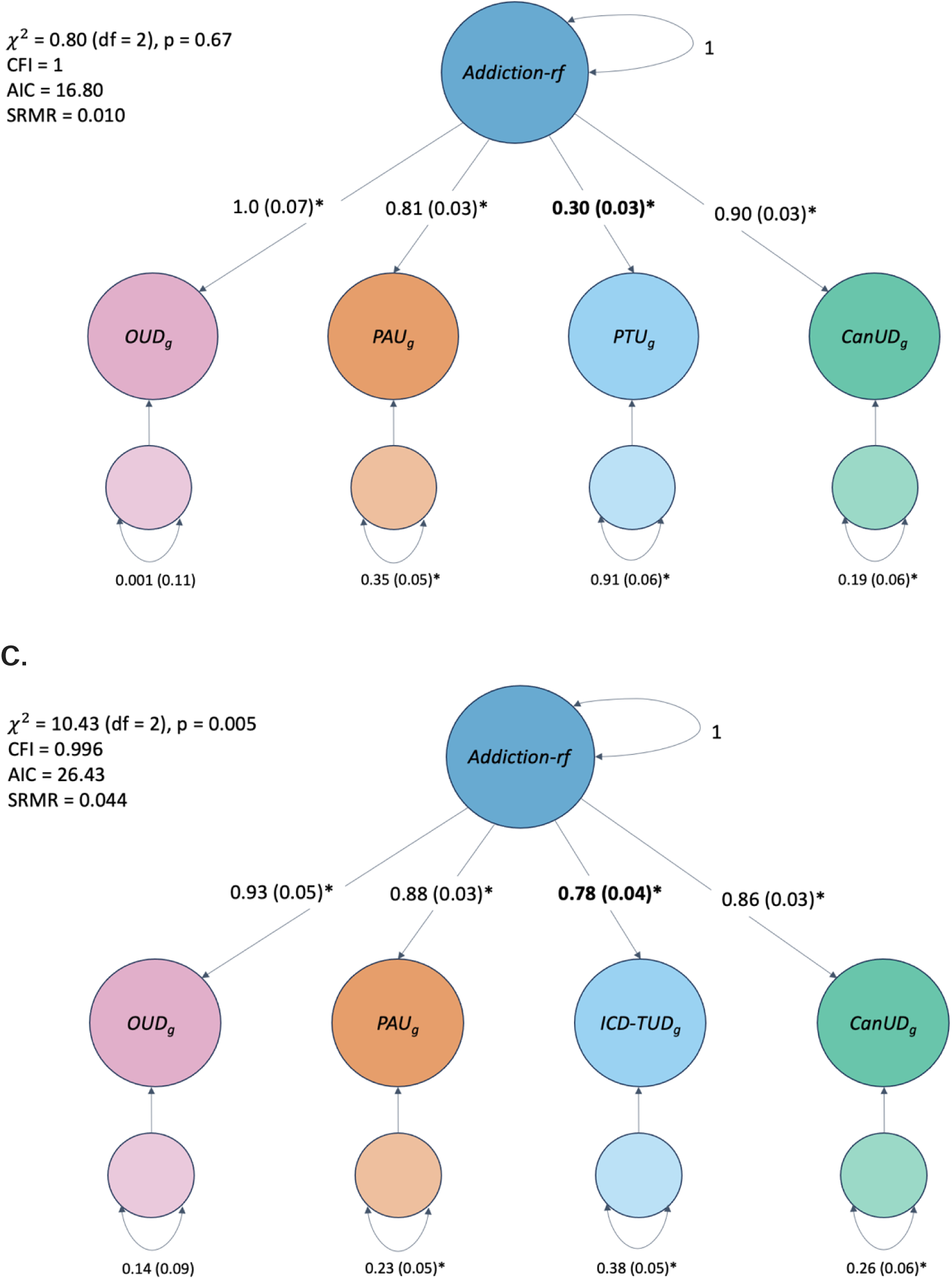
A modified Addiction-Risk-Factor model. This model is patterned upon the common factor model in Figure 1A of Hatoum *et al.*, 2022, but updated with new, larger versions of the OUD^26^, PAU^24,25^, and CanUD GWAS and using 3 different phenotypes for tobacco GWAS. **Panel A:** DSM-NicDep. **Panel B:** PTU^5^ GWAS. **Panel C:** ICD-TUD^6^. Significant loadings at p < 0.05 are represented by *. Addiction-rflJ=lJThe Addiction-Risk-Factor; OUD = opioid use disorder; PAU = problematic alcohol use; CanUD = cannabis use disorder; DSM-NicDep = nicotine dependence; ICD-TUD = ICD-based tobacco use disorder.

### Polygenic scores for DSM-NicDep and FTND

A polygenic score (PGS) for DSM-NicDep was significantly associated with DSM-5 tobacco use disorder in the European ancestry subset of the NESARC-III sample (beta = 0.07, SE = 0.02, p-value = 5.11e-3; N_cases_ = 4,205), as was a polygenic score for FTND (beta = 0.07, SE = 0.02, p-value = 3.11e-3). Results from item-level logistic regression models are shown in **Figure 3**. The DSM-NicDep PGS was associated with 6 of 11 criteria, as was the FTND PGS, although only 3 of these criteria were associated with both (TimeSpent, Problems, and Withdrawal). The FTND PGS was associated with 3 of the 4 FTND items, as expected, while the DSM-NicDep PGS was not associated with any FTND items. Failure to fulfill major role obligations (Fail) and Use of larger amounts or for longer than intended (Larger) were unrelated to both PGS after multiple testing correction. Despite these apparent disparities, estimates were not significantly different for the DSM-NicDep and FTND PGS.

**Figure 3.**
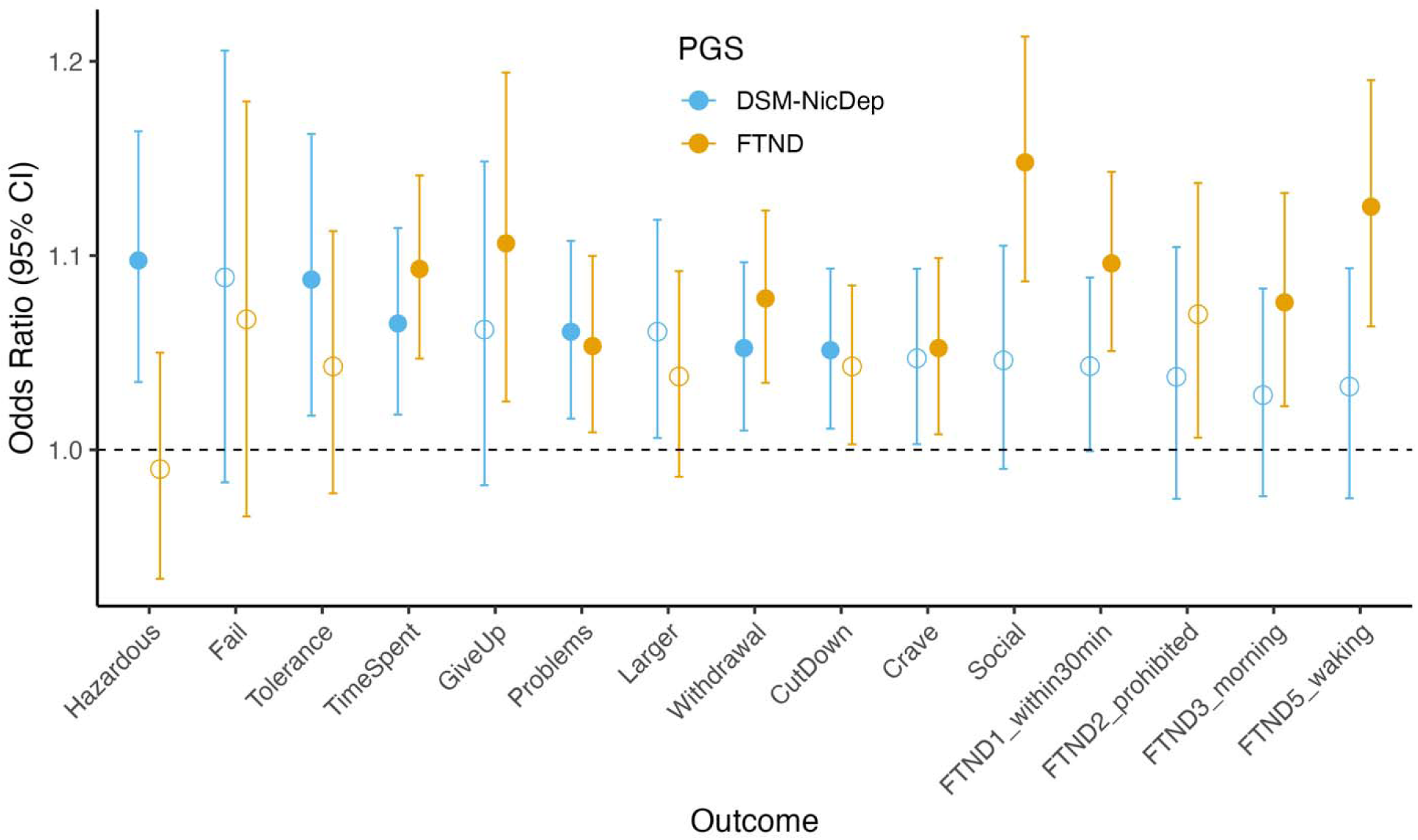
Polygenic scores (PGS) for FTND and DSM-NicDep predict individual DSM-5 nicotine use disorder and FTND criteria in the European ancestry subset of the NESARC-III sample. Filled circles represent estimates that were significant after FDR correction, while open circles represent estimates that were not significant after FDR correction. Hazardous = Recurrent use in physically hazardous situations; Fail = Recurrent use resulting in failure to fulfill major role obligations at work, school or home; Tolerance = Marked need for increased amount to get same effect, or diminished effect of same amount; TimeSpent = Great deal of time spent in activities necessary to obtain or use; GiveUp = Important recreational, social or occupational activities given up or reduced; Problems = Use despite knowledge of persistent/recurrent physical/psychological problems; Larger = Taken over larger amounts/longer periods than intended; Withdrawal = Withdrawal syndrome or use to relieve/avoid syndrome; Cutdown = Persistent desire or unsuccessful attempts to cutdown or control use; Crave = Craving, or strong urge or desire to use; Social = Persistent use despite recurring social/interpersonal problems caused or exacerbated by use; FTND1_within30min = How soon after you wake up do you smoke your first cigarette?; FTND2_prohibited = Do you find it difficult to refrain from smoking in places where it is forbidden?; FTND3_morning = Which cigarette would you hate most to give up?; FTND5_waking = Do you smoke more frequently during the first hours after waking than during the rest of the day?

## DISCUSSION

In this first cross-ancestry GWAS of DSM-NicDep (N = 61,861), we replicated the well-known *CHRNA5* risk locus. DSM-NicDep was genetically correlated with other substance use disorders, smoking-related phenotypes, and psychiatric disorders. DSM-NicDep was more closely related to the genetic underpinnings of other DSM- or ICD-defined substance use disorders than was PTU (problematic tobacco use, a phenotype that combined FTND and cigarettes per day).

ICD and DSM criteria focus on physical and psychological (and social, in DSM-5) impairment due to substance use, and are, for the most part, identical across substances. FTND, on the other hand, places greater emphasis on physiological aspects of dependence (e.g., items related to tolerance and withdrawal^12^). Because most GWAS of substance use disorders utilize ICD and DSM criteria, we hypothesized that this discrepancy might underlie the lower genetic correlation observed between nicotine dependence and other substance use disorders in prior twin and genome-wide correlation studies that used FTND^6^. Indeed, the loading for DSM-NicDep in the Addiction-Risk-Factor genomic SEM was nearly 3 times that of the loading for PTU (0.83 vs. 0.30) (**Figure 1**). Notably, the loadings for OUD and PAU also increased in our model (0.83 to 1.0, and 0.58 to 0.81, respectively), likely due to the larger sample sizes of these more recent GWAS. We also noted no significant residual correlation between OUD and PAU in our analysis.

Polygenic score (PGS) analyses of individual criteria in NESARC-III revealed an interesting pattern of associations. First, none of the FTND-specific items were correlated with the DSM-NicDep PGS, suggesting that the DSM-NicDep PGS captured genetic variance that was distinct from the FTND items. Second, even though DSM-IV did not include a diagnosis of nicotine abuse, DSM-5 includes 3 abuse criteria in its diagnosis of Tobacco Use Disorder – of these, two were unrelated to the DSM-NicDep PGS (Social, Fail) while the third (Hazard) was significantly associated. Notably, Fail was unrelated to both PGS and was amongst the least commonly endorsed criteria. Third, while the FTND is presumed to evaluate aspects of both tolerance and withdrawal, the DSM Tolerance criterion was unrelated to the FTND PGS. In contrast, Withdrawal as assessed by the DSM was associated with both PGS, while Time to first cigarette was solely associated with FTND. Taken together, these item-level analyses suggest that the DSM-NicDep GWAS may have indexed genetic liability to a distinct set of tobacco-related problems than the FTND GWAS. However, many confidence intervals on these estimates were wide. Future item-level GWAS using novel structural equation modeling methods that bring together DSM and FTND items might be of high value at parsing whether genetic and clinical heterogeneity align (**Figure 3**).

We expected that DSM-NicDep would be more genetically correlated with psychopathology than FTND, as has been found in clinical and epidemiological studies^8^. The point estimates of our genetic correlation analysis suggested that PTU (CPD + FTND) was the least associated with most indices of psychopathology, while both DSM-NicDep and ICD-TUD were more strongly associated with psychosocial indices. DSM-NicDep appeared to additionally index material deprivation more strongly than the other traits, while all four tobacco GWAS (DSM-NicDep, ICD-TUD, FTND and PTU) were equivalently related to respiratory (e.g., FEV1, Lung cancer) and metabolic markers of tobacco exposure.

The largest GWAS of CPD^3^ identified 140 genomic risk loci, compared with 5 in the largest GWAS of FTND^4^ and 1 in the current study of DSM-NicDep. This likely reflects the much larger sample size in the CPD study. Despite a similar sample size, the FTND GWAS identified more loci than the current DSM-NicDep GWAS, possibly because that study used a linear model to analyze a categorical 3-level (mild-moderate-severe) measure while our analyses relied on binary diagnoses. A recent GWAS of ICD-TUD that had a much larger sample size than ours (N = 898,680) identified 88 loci^6^. In the current study, DSM-NicDep was highly correlated with ICD-TUD. Given their high genetic correlation, future efforts may consider combining these diagnostic modalities as sources of information on tobacco use disorder.

We did not find any significant loci in the much smaller African or East Asian ancestry analyses, due to the lack of power. The lack of diversity in genetic data is unfortunate as nicotine dependence and tobacco use disorder are leading contributors to mortality in worldwide populations^2,42^. A related limitation was our inability to conduct sex- and birth cohort-stratified analyses; rates of tobacco use vary markedly according to both sex and birth cohort, which may in turn modify the extent to which genetic liability influences risk for DSM-NicDep. Amassing more genetic datasets with DSM-derived nicotine dependence would enable such valuable analyses.

In summary, our analyses highlight the importance of considering diagnostic assessment in genetic studies of substance use disorders. We found that DSM-NicDep was more closely related to a general addiction liability factor compared to a “problematic tobacco use” phenotype that combined FTND and CPD. The DSM-NicDep PGS was not significantly associated with FTND criteria in an independent sample. Compared to FTND, DSM-NicDep was more strongly genetically correlated with ICD-TUD, smoking initiation, cannabis ever-use, and material deprivation. Given the strong genetic correlation between DSM-NicDep and ICD-TUD, future analyses may consider combining data from DSM-NicDep and ICD-based studies of TUD to maximize sample size for gene discovery.

## Data Availability

Upon publication, summary statistics from this study will be made available at the following website for those who agree to follow the data access terms and conditions: https://pgc.unc.edu/for-researchers/download-results/

## Author Contributions

ECJ, DL, HJE, JG, and AA conceived the study idea. ECJ, DL, APM, ASH, JDD, JVB, MG, KS, TT, TE, PJ, TK, CM, YN, TP, MS, PNRV, LW, and IRG contributed to data analysis. ECJ, DL, and AA drafted the manuscript, and all authors edited, reviewed, and approved the final manuscript.

## Acknowledgements

We gratefully acknowledge our contributing studies and the participants in those studies without whom this effort would not be possible.

## AGDS

We thank all the participants for giving their time to contribute to this study. We wish to thank all the people who helped in the conception, implementation, beta testing, media campaign and data cleaning. We would specifically like to acknowledge Ken Kendler, Patrick Sullivan, Andrew McIntosh and Cathryn Lewis for input on the questionnaire; Lorelle Nunn, Mary Ferguson, Lucy Winkler and Natalie Garden for data and sample collection; Natalia Zmicerevska, Alissa Nichles and Candace Brennan for participant recruitment support. Jonathan Davies, Luke Lowrey and Valeriano Antonini for support with IT aspects; Vera Morgan and Ken Kirkby for help with the media campaign.

## GBP

We thank the participants for giving their time and support for this project. We acknowledge and thank M. Steffens for her generous donations in loving memory of J. Banks. Funding support for the **Comorbidity and Trauma Study (CATS)** (dbGAP accession number: phs000277.v1.p1) was provided by the National Institute on Drug Abuse (R01 DA17305); GWAS genotyping services at the CIDR at The Johns Hopkins University were supported by the National Institutes of Health (contract N01-HG-65403). The **Christchurch Health and Development Study (CHDS**: dbGAP in progress) has been supported by funding from the Health Research Council of New Zealand, the National Child Health Research Foundation (Cure Kids), the Canterbury Medical Research Foundation, the New Zealand Lottery Grants Board, the University of Otago, the Carney Centre for Pharmacogenomics, the James Hume Bequest Fund, US NIH grant MH077874, and NIDA grant ‘‘A developmental model of gene-environment interplay in SUDs’’ (R01DA024413) 2007–2012. The **Collaborative Study on the Genetics of Alcoholism (COGA)**, Principal Investigators B. Porjesz, V. Hesselbrock, A. Agrawal; Scientific Director, A. Agrawal; Translational Director, D. Dick, includes ten different centers: University of Connecticut (V. Hesselbrock); Indiana University (H.J. Edenberg, T. Foroud, Y. Liu, M.H. Plawecki); University of Iowa Carver College of Medicine (S. Kuperman, J. Kramer); SUNY Downstate Health Sciences University (B. Porjesz, J. Meyers, C. Kamarajan, A. Pandey); Washington University in St. Louis (L. Bierut, J. Rice, K. Bucholz, A. Agrawal, S. Hartz); University of California at San Diego (M. Schuckit); Rutgers University (J. Tischfield, D. Dick, R. Hart, J. Salvatore); The Children’s Hospital of Philadelphia, University of Pennsylvania (L. Almasy); Icahn School of Medicine at Mount Sinai (A. Goate, P. Slesinger); and Howard University (D. Scott). Other COGA collaborators include: L. Bauer (University of Connecticut); J. Nurnberger Jr., L. Wetherill, X., Xuei, D. Lai, S. O’Connor, (Indiana University); G. Chan (University of Iowa; University of Connecticut); D.B. Chorlian, J. Zhang, P. Barr, S. Kinreich, G. Pandey, Z. Neale (SUNY Downstate); N. Mullins (Icahn School of Medicine at Mount Sinai); A. Anokhin, S. Hartz, E. Johnson, V. McCutcheon, S. Saccone (Washington University); J. Moore, F. Aliev, Z. Pang, S. Kuo (Rutgers University); A. Merikangas (The Children’s Hospital of Philadelphia and University of Pennsylvania); H. Chin and A. Parsian are the NIAAA Staff Collaborators. We continue to be inspired by our memories of Henri Begleiter and Theodore Reich, founding PI and Co-PI of COGA, and also owe a debt of gratitude to other past organizers of COGA, including Ting-Kai Li, P. Michael Conneally, Raymond Crowe, and Wendy Reich, for their critical contributions. This national collaborative study is supported by NIH Grant U10AA008401 from the National Institute on Alcohol Abuse and Alcoholism (NIAAA) and the National Institute on Drug Abuse (NIDA). Support for the **Study of Addiction: Genetics and Environment (SAGE)** was provided through the NIH Genes, Environment and Health Initiative [GEI; U01 HG004422; dbGaP study accession phs000092.v1.p1]. SAGE is one of the genome-wide association studies funded as part of the Gene Environment Association Studies (GENEVA) under GEI. Assistance with phenotype harmonization and genotype cleaning, as well as with general study coordination, was provided by the GENEVA Coordinating Center [U01 HG004446]. Assistance with data cleaning was provided by the National Center for Biotechnology Information. Support for collection of datasets and samples was provided by the Collaborative Study on the Genetics of Alcoholism [COGA; U10 AA008401], the Collaborative Genetic Study of Nicotine Dependence [COGEND; P01 CA089392; see also phs000404.v1.p1], and the Family Study of Cocaine Dependence [FSCD; R01 DA013423, R01 DA019963]. Funding support for genotyping, which was performed at the Johns Hopkins University Center for Inherited Disease Research (CIDR), was provided by the NIH GEI [U01HG004438], the National Institute on Alcohol Abuse and Alcoholism, the National Institute on Drug Abuse, and the NIH contract “High throughput genotyping for studying the genetic contributions to human disease” [HHSN268200782096C]. The **GSMS** project (phs000852.v1.p1) was supported by the National Institute on Drug Abuse (U01DA024413, R01DA11301), the National Institute of Mental Health (R01MH063970, R01MH063671, R01MH048085, K01MH093731 and K23MH080230), NARSAD, and the William T. Grant Foundation. We are grateful to all the GSMS and CCC study participants who contributed to this work. The following grants supported data collection and analysis of **CADD** (dbGAP in progress): DA011015, DA012845, DA021913, DA021905, DA032555, and DA035804. **ADAA** was funded by NIH grant R01 AA017444. **Brisbane Longitudinal Twin Study (BLTS)** was supported by the United States National Institute on Drug Abuse (R00DA023549), and by the Australian Research Council (DP0343921, DP0664638, 464914, 619667, FT110100548). **Yale-Penn** (phs000425.v1.p1; phs000952.v1.p1) was supported by National Institutes of Health Grants RC2 DA028909, R01 DA12690, R01 DA12849, R01 DA18432, R01 AA11330, and R01 AA017535 and the Veterans Affairs Connecticut and Philadelphia Veterans Affairs Mental Illness Research, Educational, and Clinical Centers. **Australian Alcohol and Nicotine studies (OZ-ALC-NAG**; phs000181.v1.p1) were supported by National Institutes of Health Grants AA07535,AA07728, AA13320, AA13321, AA14041, AA11998, AA17688, DA12854, and DA019951; by Grants from the Australian National Health and Medical Research Council (241944, 339462, 389927,389875, 389891, 389892, 389938, 442915, 442981, 496739, 552485,and 552498); by Grants from the Australian Research Council(A7960034, A79906588, A79801419, DP0770096, DP0212016, and DP0343921); and by the 5th Framework Programme (FP-5) GenomEUtwin Project (QLG2-CT-2002-01254). Genome-wide association study genotyping at Center for Inherited Disease Research was supported by a Grant to the late Richard Todd, M.D., Ph.D., former Principal Investigator of Grant AA13320. The **Finnish Twin Cohort/Nicotine Addiction Genetics-Finland** study was supported by Academy of Finland (grants # 213506, 129680 to JK), NIH DA12854 (PAFM), Global Research Award for Nicotine Dependence / Pfizer Inc. (JK), Wellcome Trust Sanger Institute, UK and the European Community’s Seventh Framework Programme ENGAGE Consortium (HEALTH-F4-2007-201413). In **Finntwin12**, support for data collection and genotyping has come from National Institute of Alcohol Abuse and Alcoholism (grants AA-12502, AA-00145, and AA-09203 to RJR and AA15416 and K02AA018755 to DMD), the Academy of Finland (grants 100499, 205585, 118555, 141054 and 264146 to JK) & Wellcome Trust Sanger Institute, UK.

