## Supplemental Methods for "Multi-ancestral genome-wide association study of clinically defined nicotine dependence reveals strong genetic correlations with other substance use disorders and health-related traits"

***Sample descriptions***

**deCODE**

*Sample description:* Cases were drawn from the largest addiction treatment center in Iceland, the SAA‐National Center of Addiction Medicine. Controls were recruited as part of various genetic research programs at deCODE Genetics. Individuals diagnosed with substance use/abuse were excluded from controls. The deCODE Genetics study was approved by the Data Protection Commission of Iceland and the National Bioethics Committee of Iceland.

*Nicotine dependence measure:* Nicotine dependence diagnoses in this treatment cohort were made, in the years 1977 – 2014, using DSM-IIIR, DSM-IV and DSM-5 criteria, by clinicians using the Diagnostic and Statistical Manual of Mental Disorders (DSM) system.

**Brisbane Longitudinal Twin Study (BLTS)**

*Sample description:* The BLTS was initiated in 1992 to study melanocytic nevi. Subjects were recruited from primary and secondary schools in the greater Brisbane area through media appeals and word of mouth. To date, BLTS participants have completed up to six assessments at ages 12, 14, 16, 18, 26, and 30 years. Data for this study come from the fifth wave of the 19UP project, conducted when participants were approximately 26 years old (mean age = 25.7, SD = 4.3, range = 18.7-38.3). The 19UP project assessed a range of mental health and behavioral problems and their risk factors, including substance use and substance use disorders. The sample comprised 2,360 twins (not pairs) and 788 non-twin siblings (56% female, mean age = 26.1, SD = 4.1, range = 18-39).

*Nicotine dependence measure:* The 19UP project collected data on drug and substance use phenotypes, heritable traits, relationships, and personality through online surveys. Among the substance use phenotypes, the project assessed lifetime nicotine use, frequency of use, and included DSM-IV and DSM-5 item-level criteria for Nicotine Dependence. The Nicotine section was also modeled on the Fagerström Test for Nicotine Dependence (Heatherton et al., 1991), and was administered conditionally based on responses to the online survey. Only subjects who reported having initiated smoking (or current smoking status) and who had smoked 100 or more cigarettes (4 to 5 packs) in their lifetime were eligible for the computer-assisted telephone interview (CATI) nicotine dependence questions. A diagnosis of nicotine dependence was contingent upon having smoked ≥100 cigarettes lifetime and reporting two or more symptoms.

**Minnesota Center for Twin and Family Research (MCTFR)**

*Sample description:* The MCTFR has recruited three community cohorts of twin pairs; these participants have been assessed 4 to 8 times with ages of assessment ranging from 10-49 and birth years ranging from 1972-1994. Data for this study come from the CO-MN interview assessment conducted between 2018-2022.

*Nicotine dependence measure:* The interview contained items on tobacco consumption and DSM-IV nicotine dependence. All participants were asked if they had ever tried any tobacco product in their lifetime, and if so, they were asked if they “smoked cigarettes more than 20 times or used any other form of tobacco more than five times or had smoked every day for at least two weeks”. Only participants responding yes to one of these three conditions received the nicotine dependence symptom questions. A diagnosis of ND was assigned to individuals who endorsed at least three symptoms.

**Australian Genetics of Depression Study (AGDS)**

*Sample description:* The AGDS Study recruited >20,000 Australian adults aged 18-90 years old who have lived experience of depression (Byrne et al. 2020).

*Nicotine dependence measure:* Participants completed an online self-report questionnaire. Lifetime nicotine dependence was assessed on DSM-5 criteria using the Composite International Diagnostic Interview (CIDI). Controls in the current analyses had smoked ≥100 cigarettes in their lifetime and were aged 25 years or older.

**Australian Genetics of Bipolar Disorder Study (GBP)**

*Sample description:* The GBP Study is a nation-wide cohort of >5,000 Australian adults aged 18-90 years old with lived experience of bipolar disorder (BD) (Lind et al. 2023). The study aims to detect the relationships between genetic risk, symptom severity, and the lifetime prevalence of BD, treatment-response and medication side-effects, and patterns and costs of health care usage.

*Nicotine dependence measure:* Participants completed an online self-report questionnaire. Lifetime nicotine dependence was assessed on DSM-5 criteria using the Composite International Diagnostic Interview (CIDI). Controls in the current analyses had smoked ≥100 cigarettes in their lifetime and were aged 25 years or older.

**University of California San Francisco (UCSF) Family Study**

*Sample Description:* The UCSF Family Alcoholism study includes small nuclear families and unrelated participants. Study probands with a lifetime history of DSM-IV Alcohol Dependence were recruited nationwide, and after agreeing to participate, their relatives were invited by mail to participate. A total of 2,154 individuals aged 18-84 from 970 families were enrolled in this study.

*Nicotine dependence measure:* Lifetime diagnoses of Nicotine Dependence were determined according to the criteria of the Diagnostic and Statistical Manual of Mental Disorders, fourth edition (DSM-IV) as assessed by a modified version of the Semi-Structured Assessment for the Genetics of Alcoholism (SSAGA; Bucholz et al., 1994).

**Finnish Twin Cohort - Nicotine Addiction Genetics study**

The study was approved by the Ethics committee of the Hospital District of Helsinki and Uusimaa, Finland, and by the IRB of Washington University, St. Louis, Missouri, USA.

*Sample description:* The study sample was an intensively studied sub-sample from the Finnish Twin Cohort (FTC) consisting of adult twins born in 1938-1957. This sample was ascertained for smoking, so that based on earlier survey data, the twin pairs concordant for ever-smoking were identified and recruited along with their family members for the Nicotine Addiction Genetics (NAG) Finland study, as part of the international consortium funded by the NIH (Hällfors et al., 2018). Data were collected between 2001 and 2005 when participants were assessed by DNA sample collection and structured diagnostic psychiatric interviews. The analysis sample consisted of 2085 individuals with both phenotype and genotype data available (54.2 percent males, mean age 56.9 years, from 748 families

*Nicotine dependence measures:* Data provides detailed phenotypic information on multiple smoking behavior traits, including consumption of cigarettes per day (CPD), the Fagerström Test for Nicotine dependence (FTND), and the DSM-IV nicotine dependence diagnosis. Participants were interviewed using the diagnostic Semi-Structured Assessment for the Genetics of Alcoholism (SSAGA) protocol including an additional section on smoking and nicotine dependence adapted from the Composite International Diagnostic Interview (CIDI). The customized computer-assisted telephone interviews included more than 100 questions on smoking behavior. All participants provided written informed consent.

Hällfors et al. Genome-wide association study in Finnish twins highlights the connection between nicotine addiction and neurotrophin signaling pathway, Addiction Biology, 2018, 24, 549–561; doi:10.1111/adb.12618

**Comorbidity and Trauma Study (CATS)**

*Sample description:* This study consisted of opioid dependent individuals aged 18 and older recruited from opioid substitution therapy clinics in the greater Sydney area and genetically unrelated individuals with little or no lifetime opioid misuse from neighborhoods in geographic proximity to these clinics. All subjects were of European-Australian descent.

*Nicotine dependence measure:* All participants were assessed using a version of the Semi-Structured Assessment for the Genetics of Alcoholism (SSAGA). Nicotine dependence was defined using DSM-IV criteria. For the purposes of these analyses, controls were defined as anyone who reported ever smoking.

**Christchurch Health and Development study (CHDS)**

*Sample description:* The Christchurch Health and Development study (CHDS)8,9 is a

longitudinal study of a birth cohort of 1,265 children collected in mid-1977 from urban Christchurch, New Zealand. Data on social circumstances, health, development and wellbeing of the participants was obtained from the cohort at birth, four months, one year, annually to age 16 years, and at 18, 21, 25, 30, and 35 years. All study information was collected on the basis of

signed consent from study participants and all information is fully confidential. All aspects of the study have been approved by the Canterbury (NZ) Ethics Committee.

*Nicotine dependence measure*: At ages 18, 21, 25, 30 and 35 years cohort members were questioned about their substance use behaviours and problems associated with substance use since the previous assessment (alcohol, tobacco, cannabis, other illicit drugs), using the relevant sections of the Composite International Diagnostic Interview (CIDI) to assess DSM-IV symptom criteria for substance use disorders. Using this information, lifetime nicotine

dependence was classified on the basis of whether the participant met DSM criteria for nicotine dependence at any assessment up to age 35. Controls were individuals exposed to nicotine but who did not meet criteria for dependence.

**Collaborative Study on the Genetics of Alcoholism (COGA)**

*Sample description:* COGA is a multi-site study of alcohol dependent probands and their family members. Alcohol dependent probands were recruited from inpatient and outpatient facilities. Community probands and their family members were also recruited from a variety of sources. The full sample of 12,145 individuals were genotyped on four different genome-wide genotyping arrays. Among these arrays, two to 127 samples were genotyped on at least two different arrays

with pairwise concordance rates all > 99.18%.

*Nicotine dependence measure* : All participants were assessed using the Semi-Structured Assessment for the Genetics of Alcoholism. Cases met criteria for a lifetime history of DSM-IV nicotine dependence. Individuals not meeting criteria for nicotine dependence were included as controls.

**Study of Addiction: Genetics and Environment (SAGE), Collaborative Genetic Study of**

**Nicotine Dependence (COGEND) & Family Study of Cocaine Dependence (FSCD)**

*Sample description:* Subjects for the Study of Addiction: Genetics and Environment (SAGE) were selected from three large, complementary studies: COGA, Family Study of Cocaine Dependence (FSCD), and the Collaborative Genetic Study of Nicotine Dependence (COGEND). Overlapping individuals were removed across these studies. COGA participants were assessed using the Semi-Structured Assessment for the Genetics of Alcoholism (SSAGA). FSCD and COGEND participants were assessed using polydiagnostic instruments closely based on the SSAGA.

*Nicotine dependence measure:* Cases had a lifetime history of meeting criteria for DSM-5 tobacco use disorder. Controls were individuals who did not meet case criteria but did report a lifetime history of ever-smoking and were 25 or older.

**Center on Antisocial Drug Dependence (CADD)**

*Sample description:* The sample of 1,901 unrelated adolescents was aggregated from several

studies described elsewhere. This cohort was over-selected for adolescent behavioral

disinhibition, with half of the participants ascertained specifically from high-risk populations (i.e.

recruited through substance abuse treatment, special schools, or involvement with the criminal

justice system; see supplement of 20 for additional criteria for clinical probands). CADD GWAS

participants were an average age of 16.5 (SD = 1.4, range = 13.0–19.9), 28.9% were female, and 37.3% of participants reported non-Caucasian ancestry.

*Nicotine dependence measure*: Lifetime nicotine dependence was assessed with the CIDI-SAM and defined as meeting nicotine dependence at any wave for this longitudinal study.

**Gene-Environment-Development Initiative (GEDI) – Duke University (GSMS)**

*Sample description:* The Duke arm of the NIDA-funded Gene-Environment-Development Initiative (GEDI) combined existing phenotypic and environmental data from two large prospective studies, the Great Smoky Mountains Study (GSMS) and the Caring for Children in the Community (CCC) study. For each of the two population-based contributing studies, genome-wide genotyping was conducted using a common platform (Illumina Human660W- Quad v1), generating a total genotyped sample of ~1300 subjects.

*Nicotine dependence measure:* Participants of both studies were assessed via structured interviewing using the Young Adult Psychiatric Assessment and its early life extension (i.e., YAPA and CAPA), yielding diagnoses and symptom scales for a wide range of substance use disorders (SUDs). Nicotine dependence was defined using DSM-IV criteria.

**Australian Alcohol and Nicotine Studies (OZALC)**

*Sample description:* Participants were recruited from twins and their relatives who had participated in questionnaire- and interview-based studies on alcohol and nicotine use and alcohol-related events or symptoms. They were living in Australia and of predominantly European ancestry.

*Nicotine dependence measure:* Nicotine dependence was assessed using DSM-IV criteria.

**Alcohol Dependence in African Americans (ADAA)**

*Sample description:* Data from “Alcohol Dependence in African Americans: A Case-Control Genetic Study” (ADAA) was funded by NIH grant R01 AA017444. The data were collected between 2009 and 2013 and consisted of cases recruited from treatment centers in St. Louis, Missouri and controls screened for the absence of alcohol use disorder recruited from households selected from neighborhoods in proximity to neighborhoods of residence of case participants.

*Nicotine dependence measure*: Cases met criteria for DSM-5 tobacco use disorder. Controls were individuals who did not meet case criteria but did report a lifetime history of ever-smoking and were 25 or older.

**Yale Penn**

*Sample description:* Yale-Penn subjects were recruited in the eastern US, predominantly in Connecticut and Pennsylvania. They were administered the Semi-Structured Assessment for Drug Dependence and Alcoholism (SSADDA) to derive DSM-IV diagnoses of lifetime alcohol and drug dependence (and other major psychiatric traits). The study received IRB approval from all participating institutions and written informed consent was obtained from all study participants.

*Nicotine dependence measure:* DSM-IV diagnoses came from the Semi-Structured Assessment for Drug Dependence and Alcoholism (SSADDA).

**Genetics of Substance Dependence in Thailand**

*Sample description:* Data from “Genetics of Substance Dependence in Thailand” was funded by NIH grants R01DA037974, D43TW009087, D43TW006166, and D43TW012262. The data were collected between 2007 and 2023 and consisted of subjects recruited with the Chulalongkorn University Faculty of Medicine, at the Princess Mother National Institute on Drug Abuse Treatment (PMNIDAT; formerly Thanyarak Institute) located in Bangkok, and Suan Prung Psychiatric Hospital in Chiang Mai. According to the stage of their recruitment, and the array used for DNA-sampling, there are three subsamples:

- Global Screening Array (“GSA”)
- Multi-Ethnic Global Array (“MEGA”)
- Asian Screening Array (“ASA”)

*Nicotine dependence measure*: Cases met criteria for DSM-IV tobacco use disorder. Controls were individuals who did not meet case criteria but did report a lifetime history of ever-smoking.

***GWAS included in genetic correlation analyses***

| **Trait** | **Study PMID** |
| --- | --- |
| FTND | 33144568 |
| PTU | 34750568 |
| ICD-TUD | 38632388 |
| CanUD | 37985822 |
| OUD | 35879402 |
| PAU | 38062264 |
| CanUse | 30150663 |
| DPW | 36477530 |
| CPD | 36477530 |
| SmkCessation | 36477530 |
| SmkInit | 36477530 |
| ADHD | 36702997 |
| Anxiety | 31906708 |
| Depression | 34045744 |
| PTSD | 31594949 |
| Schizophrenia | 35396580 |
| Suicide attempt | 34861974 |
| Cot+HC | 32157176 |
| FEV1 | http://www.nealelab.is/uk-biobank/ |
| Lung cancer | 28604730 |
| Risk tolerance | 30643258 |
| Edu attainment | 30038396 |
| EF | 36150907 |
| TDI | http://www.nealelab.is/uk-biobank/ |
| BMI | 30124842 |
| Height | 36224396 |

CanUD = cannabis use disorder; OUD = opioid use disorder; PAU = problematic alcohol use, ICD-TUD = ICD-based tobacco use disorder; CanUse = cannabis ever-use; DPW = drinks per week; SmkInit = smoking initiation, SmkCessation = smoking cessation, CPD = cigarettes per day; ADHD = attention deficit hyperactivity disorder; PTSD = post-traumatic stress disorder; Cot+HC = cotinine + 3-hydroxycotinine; FEV1 = forced expiratory volume in 1 second; Edu attainment = educational attainment; TDI = Townsend deprivation index; EF = executive function; BMI = body mass index.

Note: we attempted to also estimate the genetic correlation between DSM-NicDep and the nicotine metabolite ratio (NMR) from PMID 32157176, but LDSC was unable to estimate the genetic correlation, likely because of the small sample size of the NMR GWAS.

***NESARC-III Sample Description, Genetic QC, & Imputation***

The National Epidemiologic Survey on Alcohol and Related Conditions-III (NESARC-III) sample is a large, representative community survey on alcohol and other substance use behaviors in the United States, with oversampling of African American and Hispanic participants. The NESARC-III sample includes diagnostic and item level data on 22,848 individuals. The sample was genotyped using the Affymetrix Axiom® Exome Array, consisting of a total of 422,687 SNPs, and the European ancestry subset of the sample were imputed to the 1000 Genomes European ancestry reference panel using the Ricopili pipeline.
